# Avoiding COVID-19: Aerosol Guidelines

**DOI:** 10.1101/2020.05.21.20108894

**Authors:** Matthew J. Evans

**Affiliations:** Department of Physics Massachusetts Institute of Technology Cambridge, MA 02139

**Keywords:** SARS-CoV-2, COVID-19, Coronavirus, Aerosol, airborne

## Abstract

The COVID-19 pandemic has brought into sharp focus the need to understand respiratory virus transmission mechanisms. In preparation for an anticipated influenza pandemic, a substantial body of literature has developed over the last few decades showing that the short-range aerosol route is an important, though often neglected transmission path. We develop a simple mathematical model for COVID-19 transmission via aerosols, apply it to known outbreaks, and present quantitative guidelines for ventilation and occupancy in the workplace.

## 1 Introduction

The world is learning to navigate the COVID-19 pandemic and a great deal of information about the disease is already available [1, 2, 3]. In order to adjust to this new reality it is important to understand what can be done to avoid infection, and to avoid infecting others.

SARS-CoV-2, the coronavirus which causes COVID-19, is thought to be transmitted via droplets, surface contamination and aerosols [4, 5, 6, 7, 8, 9, 10]. COVID-19 outbreaks are most common in indoor spaces [11], and evidence points to a dominant role of aerosol transmission [12, 13]. Furthermore, infection via inhalation of aerosols and small droplets dominates large droplet exposure in most cases for physical reasons linked to complex, but well understood fluid dynamics [14, 15, 16, 17]. Despite this fact, there is a common misconception that aerosol transmission implies efficient long-range transmission (as in measles), and thus that the absence of long-range transmission implies an absence of aerosol transmission. The truth is more nuanced, and includes the possibility of a dominant short-range aerosol path limited by pathogen dilution, deposition, and decay [18, 19, 20].

The production of aerosolized viruses by a contagious individual occurs naturally as a result of discrete events (e.g., coughing or sneezing), or through a continuous process like breathing and talking [21, 22]. Droplets that are formed with diameters less than ~ 50 *μ*m quickly lose most of their water to evaporation, shrinking by a factor of 2-3 in diameter and becoming “droplet nuclei”. These fine particles settle slowly and mix with the air in the environment [23, 24, 25].

Aerosol transmission happens when a susceptible individual inhales these now sub-20 *μ*m droplet nuclei that are suspended in the air around them [26, 27]. This is thought to be the dominant transmission mechanism for influenza and rhinovirus [26, 28] and possibly also for SARS and MERS [29]. For influenza, it has been shown that aerosol particles as small as 1.5 *μ*m are sufficient for transmission [30]. Furthermore, it is known from other viral respiratory diseases that aerosol exposure can result in infection and illness at much lower doses than other means (e.g., nasal inoculation) [31].

In line with current recommendations [32], we will assume that hand-hygiene protocols are being followed such that surface contamination remains a sub-dominant transmission route [12, 9]. We will also assume that contagious individuals are wearing some kind of face covering which is sufficient to disrupt the momentum of any outgoing airflow [33, 34] and catch large droplets [35]. These actions ensure that the aerosols investigated here remain the dominant transmission mechanism.

In the following sections we present a simple model for aerosol transmission in a variety of contexts, apply this model to known outbreaks, and develop guidelines for reducing the probability of transmission in the workplace.

## 2 Risk Assessment

In this section we motivate the analysis that follows as a foundation for guidelines based on the the probability that a contagious individual will infect a coworker, *P*_inf_, and the absolute risk of infection during an epidemic such as COVID-19. The later of these is not addressed directly by the analysis presented in this work and depends on the prevalence of highly-contagious (and presumably asymptomatic or pre-symptomatic) individuals in the population [36, 37, 38, 39, 40]. In the context of the current COVID-19 pandemic, a return to work seems improbable if more than 1 in 1000 people are unwittingly contagious on any given day. This suggests that the *absolute probability* of infection should be less than *P*_inf_/1000, since our calculations of *P*_inf_ *assumes the presence of a contagious individual*. An acceptable workplace transmission risk should be chosen such that a contagious employee has only a modest chance of infecting another employee during their pre-symptomatic period, and such that the absolute risk of infection remains acceptably low (e.g., less than 0.01% if *P*_inf_ ≾ 10%).

A more quantifiable and meaningful approach to risk assessment can be derived from the relative risk of infection by COVID-19 in the workplace vs. other locations, and the associated contribution of the workplace to the growth or decline of the epidemic (i.e., the reproduction number *R*_0_) [41]. The grander objective of ensuring that the workplace’s contribution to the epidemic’s *R*_0_ is small is sufficient to ensure that the relative risk of contracting COVID-19 in the workplace is small compared with other activities (e.g., homelife or recreation). A workplace transmission probability of 10%, for instance, would contribute 0.1 to *R*_0_, meaning that for a decaying epidemic with 0.5 < *R*_0_ < 1, an employee would be 5 to 10 times more likely to be infected away from work than at work.

This relative view of risk assessment requires a somewhat counter-intuitive approach to setting guidelines: the focus should not be on limiting the probability that an employee becomes infected, but rather on limiting the quantity of pathogens *delivered to others* by a contagious and as-yet-asymptomatic employee.

## 3 Infection Model

The probability of developing an infection *P*_inf_ given exposure to a volume *V* of saliva with a concentration *ρ* of virus particles (“virions”) is

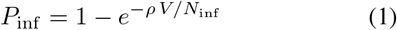

where the infectivity *N*_inf_ is the number of virions needed to make infection likely in the lungs for aerosol inhalation [21, 31, 42, 43, 20]. The infectivity value *N*_inf_ includes probable deposition location (i.e., upper vs. lower respiratory tract) and local deposition efficiency (e.g., not all inhaled droplets are deposited in the lungs [31, 44, 45]). It should be noted, however, that small doses are less likely to cause *illness* than what is indicated by Eqn. 1 [31, 46]. Rather than work explicitly with the probability of infection *P*_inf_, we will use the ratio of the viral dose and the infectivity

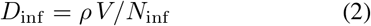

as a proxy, and refer to it as the “infectious dose”. Note that the infectivity and the viral concentration always appear together here, so only their ratio *ρ*/*N*_inf_ is important in our model. While ρ has been measured for COVID-19 [37, 47], *N*_inf_ has not been directly measured. We estimate *N*_inf_ ~ 1000 from influenza and other coronaviruses [42, 48], and check this value by applying our model to known outbreaks (see appendix A).

The relationship between *D*_inf_ and the probability of infection *P*_inf_ is shown in Fig. 1 (right). Equation 1 is shown as the dashed-grey curve, indicating that the probability of infection is high for any infectious dose greater than 1, and *P*_inf_ is essentially proportional to *D*_inf_ when *D*_inf_ ≪ 1. The other curves in Fig. 1 account for variability in *ρ*, *V* and *N*_inf_, as described in section 7.

**Figure 1:**
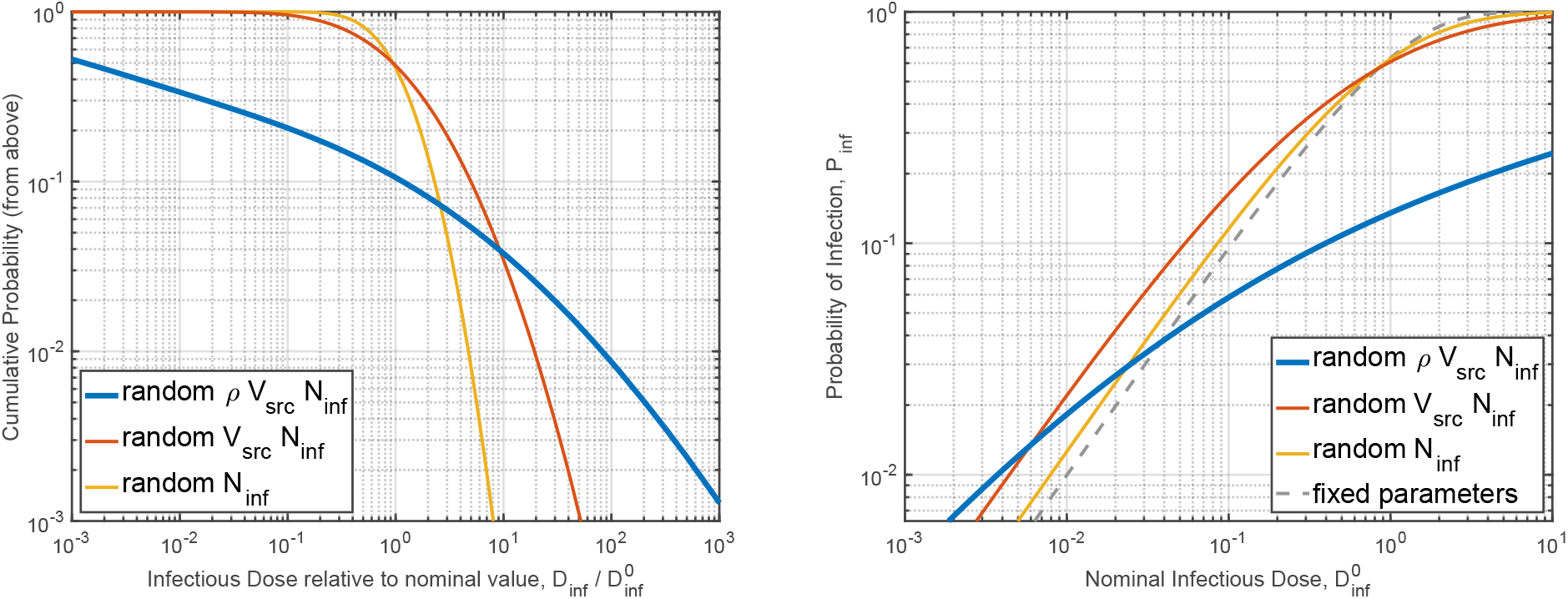
**Left**: Probability that the infectious dose in any given encounter exceeds the nominal dose by some factor. The three curves allow some or all transmission parameters to vary randomly (see section 6 and appendix B). For instance, allowing all parameters to vary (blue trace) the probability of having 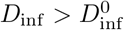 is about 10%, *D*_inf_ > 10 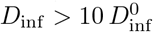 will happen in about 4% of cases, while 1% of cases will have 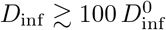. Choosing the nominal viral load *ρ*_0_ equal to the median value of 1 /nL (rather than the 90*^th^* percentile as we have) would shift the blue curve to the right by a factor of 1000. **Right**: Probability of infection as a function of nominal infectious dose 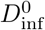. The three solid curves relate *P*_inf_ to 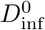 allowing some or all transmission parameters to vary randomly, while the dashed curve (grey) shows *P*_inf_ with fixed parameters as in Eqn. 1. Allowing all parameters to vary, the probability of infection for a nominal dose of 1 is 14%. 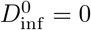 reduces *P*_inf_ to 6%, and 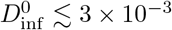 is required for *P*_inf_ < 1%. Choosing the nominal viral load *ρ*_0_ equal to 1 /nL rather than 1000 /nL would reduce 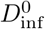 by 1000 relative to our calculations in section 4 and shift the blue curve to the left by 1000, such that *P*_inf_ would remain unchanged. The “random *V*_src_*N*_inf_” curve is used in appendix A to evaluate *P*_inf_ in cases where *ρ* has a fixed value.

## 4 Aerosol infectious Dose Model

In this section we compute the infectious dose which results from the presence of a contagious individual in various scenarios. This informs prescriptions for the maximum time a susceptible individual may be exposed to potentially contaminated air, or the minimum time between occupants in the same space. Each of these calculations uses the values in Table 1 and concludes with an exposure time limit for a given infectious dose. The infectious dose limits used here are chosen to maintain a transmission probability less than 10%, as motivated in section 2 with examples in section 7.

**Table 1:** Parameters used in the aerosol transmission model. See section 6 for more information.

| Description | Symbol | Value | Reference |
| --- | --- | --- | --- |
| Viral Load in Saliva | $\rho_0$ | 1000 /nL | [37, 47, 49] |
| Sneeze Aerosol Volume | $V_s$ | 1 μL | [50, 51] |
| Cough Aerosol Volume | $V_c$ | 100 nL | [50, 51] |
| Talking Aerosol Volume | $V_t$ | 10 nL/min | [50, 51] |
| Breathing Aerosol Volume | $V_b$ | 1 nL/min | [50, 51] |
| Aerosol Decay Time | $\tau_a$ | $\sim 20$ min | see Sec. 6.2 |
| Breathing Rate | $r_b$ | 10 L/min | [52] |
| Respiratory Infectivity | $N_{\text{inf}}$ | 1000 | [42, 31] |
| Frequently Used Symbols |  | Units | Equation |
| Infectious Dose (general) | $D_{\text{inf}}$ | number | Eqn. 2 |
| Time | $t$ | min | Eqn. 3 |
| Aerosol Source Rate | $r_{\text{src}}$ | nL/min | Eqn. 3 |
| Aerosol Source Volume | $V_{\text{src}}$ | nL | Eqn. 16 |
| Room Volume | $V_{\text{room}}$ | $\text{m}^3$ | Eqn. 4 |
| Room Ventilation Rate | $r_{\text{room}}$ | $\text{m}^3/\text{min}$ | Eqn. 4 |
| Room Air-Cycle Time | $\tau_{\text{room}}$ | min | Eqn. 4 |
| Aerosol Decay Factor | $f_a$ | number | Eqn. 7 |
| Conversion Factors |  |  |  |
| Cubic Feet per Minute | CFM | $1 \text{ m}^3/\text{min} \simeq 35 \text{ CFM}$ | |
| Air Changes per Hour | ACH | $\tau_{\text{room}} \simeq 60 \text{ min/ACH}$ | |
| Maximum Exposure and Minimum Wait Times |  |  |  |
| Room Steady State | $t_{\text{max}}^{\text{Room}} \simeq 100 \text{ min}$ | $\frac{1 \text{ nL/min}}{r_{\text{src}}} \frac{r_{\text{room}}}{10 \text{ m}^3/\text{min}}$ | Eqn. 11 |
| System Steady State | $t_{\text{max}}^{\text{Sys}} \simeq 5 \text{ hour}$ | $\frac{0.1}{1-f_{\text{sys}}} \frac{3 \text{ nL/min}}{r_{\text{src}}} \frac{f_{\text{sys}} r_{\text{sys}}}{100 \text{ m}^3/\text{min}}$ | Eqn. 14 |
| Occupancy Wait Time | $t_{\text{min}}^{\text{TE}} \simeq \tau_{\text{room}} \ln \left( 10 \frac{V_{\text{src}}}{100 \text{ nL}} \frac{1 \text{ m}^3/\text{min}}{r_{\text{room}}} \right)$ | | Eqn. 18 |
| Passage Wait Time | $t_{\text{min}}^{\text{PS}} \simeq \tau_{\text{room}} \ln \left( 10 \frac{V_{\text{src}}}{100 \text{ nL}} \frac{10 \text{ m}^3}{V_{\text{room}}} \frac{\Delta t}{1 \text{ min}} \right)$ | | Eqn. 20 |

### 4.1 Steady State in a Room

A contagious individual will shed virions into the room they occupy through breathing, talking, coughing, etc. The aerosolized virion concentration in a room can be expressed in the form of a differential equation as

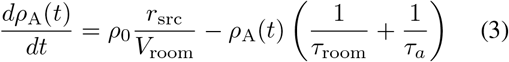

where *ρ*_0_ is the nominal viral concentration in saliva, which is emitted in aerosol form at a rate of *r*_src_, and *τ_a_* is the timescale for aerosol concentration decay. The air-cycle time in a room is

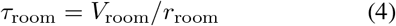

where *r*_room_ is rate at which air is removed from the room (or filtered locally). The steady-state concentration is

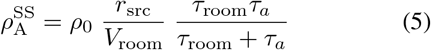

and has units of viral copies per liter of air.

Some simplification can be achieved by using Eqn. 4 to rewrite Eqn. 5 aswhere

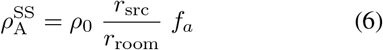

where

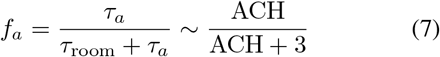

is the aerosol concentration decay factor which results from virion decay and setting of droplet nuclei (see section 6.2). The approximation gives *f_a_* in terms of air-changes per hour (ACH) for a 3 meter ceiling height. In the well ventilated limit (i.e., *τ*_room_ ≪ *τ_a_*) *f_a_* goes to 1, meaning that settling is not relevant. In poorly ventilated spaces (i.e., *τ*_room_ ≿ *τ_a_*), however, settling can play an important role since it effectively adds 3 air-changes per hour to the rate of aerosol concentration decay.

Breathing air in a room with an aerosol virion concentration *ρ_A_* will cause an accumulation of exposure *N_A_* (number of virions) proportional to the time passed in that room *t*

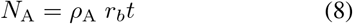

such that the steady-state infectious dose is

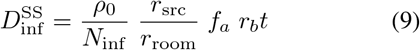

where *r_b_* ~ 10 L/min is the breathing rate. If the room is well ventilated (i.e., *f_a_* ~ 1) the infectious dose is

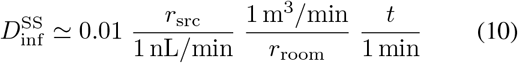

for any space (e.g. office, laboratory or bathroom) occupied by a contagious individual.

The infectious dose crosses our example event exposure threshold of 0.1 in an office-like space (*r*_room_ ~ 10 m^3^/min) after 100 minutes,

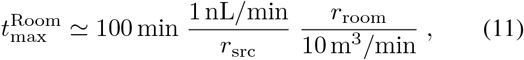

while in small spaces with less ventilation (e.g., a car or bathroom with *r*_room_ ~ 1m^3^/min) the dose would cross the threshold after only 10 minutes. This assumes that the contagious individual is quietly working such that *r*_src_ = *V_b_*, but if they are talking or coughing occasionally the source term may be much higher (e.g., *r*_src_ ≫ *V_b_*). The implication of this example is that in order to share a space for 8 hours while maintaining 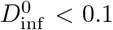, *r*_room_ should be greater than 50 m^3^/min ~ 2000 CFM per occupant beyond the first, making shared occupancy of typical office spaces untenable. Allowing for a higher nominal dose (e.g., 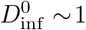), which may be acceptable if the incidence of COVID-19 in the population is low, would reduce this requirement by a factor of 10.

### 4.2 Steady State in a Building

When a susceptible individual occupies a room that is connected via the HVAC system to a room occupied by a contagious individual, there is the possibility of aerosol transmission through the HVAC system [7, 53, 54].

However, the volume of a droplet is proportional to its diameter *cubed*, which leads to the majority of the virions being delivered in the ~ 10 |im droplet nuclei [23, 30], which are filtered by HVAC systems. Most HVAC filters will remove the majority of the viral load associated with COVID-19 transmission, and a high quality filter (i.e., MERV 12) will remove at least 90% of relevant droplet nuclei [55]. The remaining aerosols are further diluted by fresh make-up air, and then spread among all of the air spaces associated with the HVAC system. The associated infectious dose in rooms connected to a room occupied by a contagious individual will be roughly

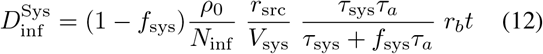

where *f*_sys_ is the fraction of the virions which is removed by the filters or displaced by make-up air, and *τ*_sys_ = *τ*_sys_/*V*_sys_ is the time required to cycle the entire air volume through the HVAC system.

In the well ventilated limit, as above, the infectious dose is

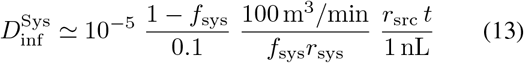

and the maximum occupancy time for 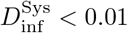 is

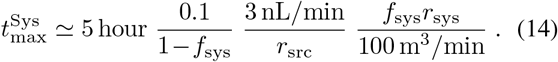

We have set the *r*_src_ scale at 3 nL/min to allow for occasional conversation (e.g., video conferencing). The maximum dose used in this example is 0.01 instead of 0.1 to allow for 10 exposed individuals (see section 7).

To understand why transmission through HVAC systems appears to be rare we note that even medium quality air filters (MERV 9) provide better than 99% filtering after some loading [55]. The value we use here assumes little or no fresh make-up air and is representative of an unloaded (i.e., clean) air filter, which is the *worst case*. Also, in buildings where the HVAC system does not recirculate air this type of transmission cannot occur (i.e., *f*_sys_ = 1).

As an explicit example, we consider 10 offices that are connected to a HVAC system which moves *r*_sys_ = 100m^3^/min with 30% make-up air and a filter that removes 90% of the virions

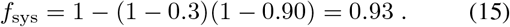

We assume that one of these offices is occupied by a contagious individual who is emitting *r*_src_ = 3 nL/min of saliva in small droplets which evaporate to form airborne droplet nuclei. According to Eqn. 13, each of the individuals in the other offices will receive an infectious dose of 0.01 after working for 8 hours. The total dose, summed over all susceptible individuals and assuming that all offices are occupied, is 0.1, indicating that there is a 6% chance that at least one of these individuals will be infected (see Fig. 1, right). Generalizing this example, we see that 10 m^3^/min ~ 350 CFM per occupant, averaged over all rooms connected by a well filtered HVAC system, is sufficient to prevent a large nominal dose.

### 4.3 Transient Occupancy and Events

Some spaces are occupied briefly by many people (e.g., bathrooms) and may not have time to come to steady state. Coughing and sneezing events can also cause a transient increase in the viral concentration in a room. The infectious dose associated with transients can be quantified as

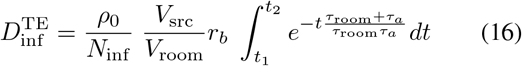

where the transient event occurs at time *t* = 0, exposure is from time *t*_1_ to *t*_2_, and *V*_src_ is the aerosol source volume (e.g., *V_c_* for a cough, or *V_s_* for a sneeze).

We can estimate when a room is “safe” for a new occupant by computing the dose in the limit of *t*_2_ → ∞,

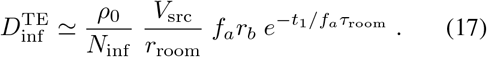

This leads to a minimum wait time of

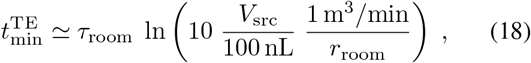

where we have set the *V*_src_ scale at 100 nL to allow for some coughing from the previous occupant, and assumed good ventilation such that *f_a_* ~ 1.

As a concrete example, for a 70 square-foot bathroom with a 70 CFM fan in operation (i.e., *τ*_room_ ≃ 10 minutes and *r*_room_ = 2m^3^/min), Eqn. 18 gives a 16 minute wait time between occupants. Increasing the airflow to 10 m^3^/min ~ 350 CFM eliminates the wait time.

Going back to Eqn. 16, we can also compute the exposure due to brief passage through a common space in which ∆*t* = *t*_2_ − *t*_1_ ≪ *τ*_room_ (e.g., corridors and elevators),

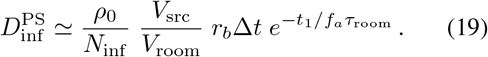

The minimum time between occupants for 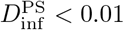 is

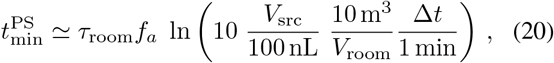

which will give negative values for large spaces or short passage times, indicating that no wait is necessary. However, this assumes well mixed air, so some mixing time is necessary to avoid a “close encounter” as described in the next section. For small spaces with poor circulation long wait times are required: a sneeze in an elevator (*V*_src_ < 1, *μ*L, *V*_room_ ~ 10 m^3^, ∆*t* ~ 1 min) would put it out-of-order for 4.6 air changes.

A stairwell, for instance, with *V*_room_ = 100 m^3^ and very little airflow (*τ*_room_ = 60 min and *f_a_* = 1/4), would require a 34 minute wait between each use to allow airflow and droplet-nuclei settling to clear out the previous occupant’s sneeze. No waiting would be required if the previous occupant was just breathing or talking (even loudly) while in the stairwell (i.e., *V*_src_ < 100 nL). However, stairwells require extra exertion during use, which will increase *r_b_*, suggesting that at least 10–15 minutes may be advisable before climbing many floors. Note that while stairwells have unusually high ceilings, which could impact the time required for droplet nuclei to settle, the ratio of volume to horizontal surface area is similar to other spaces (see section 6.2 for more discussion of settling times).

### 4.4 Close Encounters

Mixing of aerosols into an airspace may require a few air-cycle times before a fairly uniform concentration can be assumed [56, 57]. In order to understand the potential infection risk associated with close proximity, we make a very rough estimate of the exposure in the vicinity of a mask-wearing cougher. The cough momentum will be disrupted by the mask, but aerosols will exit the mask on essentially all sides [34, 33, 22] resulting in a cloud around the cougher that will then either move with local air currents or rise with the cougher’s body plume [58]. We assume that large droplets are trapped in the cougher’s mask or settle out around the cougher, while aerosols are dispersed into the air around the cougher. The infectious dose at a distance *d* for isotropic dispersion is

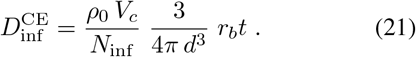

For a typical cough this leads to

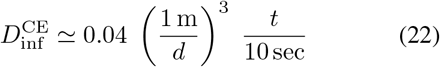

which suggests that an infection risk is still present at short distances, even if the cougher is wearing a mask [59]. This analysis is clearly oversimplified, as details of the cough, the mask, and local air currents will all prevent isotropic dispersion, but serves to emphasize that extra care should be taken for transient events at short distances as they may result in a significant infectious dose.

## 5 Mitigation Measures

The calculations in the previous section allow a variety of possible mitigation measures. This section briefly describes some means of reducing the probability of aerosol infection.

### 5.1 Masks and Respirators

Improvised face coverings, surgical masks and respirators, all collectively referred to as “masks” herein, serve a variety of purposes related to respiratory virus transmission:

1. limiting direct transmission in face-to-face interactions
2. protecting the wearer by filtering inhaled air
3. protecting others by filtering exhaled air

The first of these is the easiest to achieve: by disrupting any high-velocity outgoing pathogen-laden air flow [34, 33, 60, 14, 17] and the capturing large droplets [35, 61], masks of essentially any sort can limit direct transmission due to coughs, sneezes and conversations. This paper assumes that this goal is achieved by universal mask use, thereby allowing for our simplified analysis of well-mixed aerosols.

Masks and respirators can reduce the probability of infection, both for the wearer and for the people they interact with, by filtering the air inhaled and exhaled by the wearer [41]. While improvised face coverings generally do not remove more than half of aerosolized particles on inhalation (protection factor PF ≾ 2) [61], surgical masks provide some level of protection (PF of 2–10, typically around 3) [62, 63]. Respirators which seal against the face are far more effective protection for the wearer (e.g., PF of 8–80 for N95s) [64], but benefiting from them requires proper fit and user compliance [65, 66, 67, 68, 69, 70], which suggests that widespread use is likely to be ineffective with currently available respirators [71, 72, 68]

Filtration of outgoing air is not well tested for most kinds of masks, but there is evidence of some protective value [61, 34]. Droplets produced while breathing, talking and coughing can be significantly reduced by mask usage [73, 74, 75, 22], but much of the air expelled while coughing and sneezing goes around the mask [34].

Since effective filtration is difficult to achieve, both for incoming and outgoing air, we assign no protective value to the wearer for mask use, and assume no reduction in out going aerosols. These are conservative assumptions which can be adjusted by reducing the infectious dose according to any assumed protection factor. For example, a well fit N95 respirator should reduce the infectious dose received by the wearer by a factor of 20, and, in the absence of an exhale valve, may reduce the quantity of low-velocity outgoing aerosols by a similar factor.

### 5.2 HVAC and Portable Air Filters

Increasing air exchange rates in an HVAC system, and avoiding recirculation can both be used to reduce aerosol concentrations indoors. Consistent use of local ventilation (e.g., bathroom fans) can also help to avoid infection. In buildings and spaces where these measures are not available or not sufficient, local air filters (a.k.a., air scrubbers) can be used to increase the introduction of clean air into the space. Small stand-alone units which filter 10 m^3^/min or more are readily available and could help to increase safety in bathrooms and small offices.

As noted in sections 4.2 and 6, the particle size of interest is greater than 1 *μ*m, so special filtering technology is *not* required [55]. Care should be taken, however, when changing filters both in stand-alone units and HVAC systems as they may contain significant viral load. Stand-alone units should be disabled for at least 3 days before changing the filter to allow time for viral infectivity to decay [76]. Building HVAC filters should be changed with proper personal protective equipment, as recommended by the CDC [55].

### 5.3 Clean Rooms and HEPA Filters

Many laboratory spaces are outfitted with HEPA filters to provide clean air for laboratory operations. Clean rooms offer a clear advantage over other spaces as they are designed to provide air which is free of small particles.

If the HEPA filters are part of a recirculating air cycle, Eqn. 14 can be used with *f*_sys_ ≿ 0.999, allowing for essentially indefinite exposure times. It should be noted, however, that HEPA filters require lower air-speeds than those offered by typical HVAC systems and as such are not a “drop in replacement” option.

Laboratories that use portable clean rooms in large spaces can be treated in a similar manner, using Eqn. 13 for the air inside the clean room and Eqn. 10 to represent the clean air supplied to the laboratory space outside the portable clean room.

Any clean room environment is likely to provide sufficient air flow to make aerosol infection very unlikely in the well-mixed approximation used in section 4. This will leave close encounters, as described in section 4.4, as the dominant infection path. If there are multiple occupants in a clean room they should avoid standing close to each other, or being “down wind” of each other [77].

### 5.4 UV-C Lighting

Illumination of the air-space in patient rooms with ultra-violet light has been shown to dramatically reduce viral longevity in aerosol form, and thereby prevent infection [19, 78, 79]. This could be used to decrease *τ_a_* in spaces where increasing air circulation (i.e., reducing *τ*_room_) is impractical. Clearly, this requires that air in the space pass through the UV-C illuminated volume, and that it received a long enough exposure to make inactivation effective. Care is also required to avoid exposing occupants to UV-C radiation, which is required for viral deactivation but can be harmful to the eyes and skin. This could be done geometrically (e.g., only illuminate spaces above 2.5 m), or actively with motion sensors.

## 6 Model Parameters

The parameters used in this model are presented in Table 1. All of these parameters vary between individuals and events, and as such the values given here are intended to serve as a means of making rough estimates which can guide decision making. This section describes the provenance of these values and their variability.

### 6.1 Viral Shedding

The volume of saliva produced in a variety of activities is used to understand the emission of virions into the environment (known as “viral shedding”). The values given here are for “typical” individuals and behaviors, and actual values for any given person or event may vary by an order of magnitude [22, 50, 51, 80, 81, 24]. Droplets produced while speaking, for instance, depend on speech loudness; speaking loudly, yelling or singing can produce an order of magnitude more saliva than speaking normally [25]. We are careful to avoid quantifying saliva production in terms of the number of droplets produced, since the large droplet-nuclei are relatively small in number but carry most of the virions (i.e., slope of the number distribution is too shallow to compensate the fact that volume goes with diameter cubed) [50, 82]. For a viral load of 1000 /nL, for example, a 1 *μ*m droplet nucleus has only a 1% probability of containing a single virion, while a 10 *μ*m droplet nucleus is likely to contain 5–15 virions [25].

The viral load in saliva, *ρ*, has been seen to vary by more than 8 orders of magnitude in individuals that test positive for COVID-19 [37, 47]. Roughly 90% of individuals tested have a viral load less than *ρ*_0_ = 1000 /nL, while 1% may have a viral load above 3 × 10^4^ /nL [47]. Viral load is seen to decline after symptom onset, so the pre-symptomatic viral load may be on the high-end of the distribution [37, 49, 83]. We use *ρ*_0_ = 1000 /nL for our nominal dose calculations because it results in a rough estimate of the probability of infection relative to the full distribution for probabilities of a few percent (i.e., the “random *ρV*_src_*N*_inf_” curve is close to the other curves around *D*_inf_ ~0.03 in Fig. 1, right). This choice does not impact the final probability of infection shown in Fig. 1, since a different choice of *ρ*_0_ would shift the “random *ρV*_src_*N*_inf_” curve to compensate.

### 6.2 Aerosol Decay Time

Ventilation rates aside, there are two timescales relevant to the decay of the infectious aerosol concentration: the decline in potency or “infectivity” of virions over time, and the slow settling of droplet nuclei due to gravity. The infectivity decay time of SARS-CoV-2 in aerosol form has been found to be *τ*_decay_ ≿ 90 min [76, 84]. Note that we are using the 1/*e* decay time, not the half-life, for ease of computation.

The second timescale is set by the settling time of the larger aerosol particles which contain the majority of the virions shed into the environment. Using the “continuous fallout” model presented in [14] and the data presented in [24, 25] we estimate this as

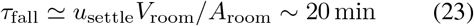

for a room with a 3 m ceiling (i.e., the ratio of the room volume to the floor area *A*_room_ is 3 m). We use *u*_settle_ ~ 0.1 m/min ~ 1 mm/s for the characteristic settling speed of speech-generated droplet nuclei [25, 24, 85], corresponding to a particle with an aerodynamic diameter of approximately 5 *μ*m.

Since the decay of the infectious aerosol concentration is dominated by settling rather the infectivity decay of the virions themselves, we use *τ_a_* ~ 20 min. The impact of settling is non-negligible in poorly ventilated spaces, as demonstrated by the outbreak described in appendix A.1.

### 6.3 Viral Intake and Infection

The rate of air exchange with the environment while breathing (“breathing rate” *r_b_*) varies among individuals and activities by a factor of a few, except in the case of strenuous exercise which can increase it by as much as a factor of 5 [21]. As such, this is a relatively well determined parameter and we use only the nominal value in our calculations. Our nominal value, *r_b_* = 10L/min, corresponds to a standing individual who is resting or engaged in light exercise, such that their tidal volume is about 0. 5 L, and their breathing cycle has a 3 s period [52].

The respiratory infectivity, *N*_inf_, is not well known for SARS-CoV-2, but it has been measured for SARS [42], other coronaviruses, and influenza [31, 48]. Variation by an order of magnitude among individuals is observed both for coronaviruses and influenza. The number we use, *N*_inf_ = 1000, is intended to be “typical” for coronaviruses [86] and represents roughly 100 “plaque forming units” (PFU) as measure for SARS [42], each of which is roughly 10 virions [87].

## 7 Transmission Probability

In this section we relate the infectious dose calculations presented in section 4 to the probability of infection (for a single individual) or transmission (to any member of a group) while accounting for the probability distributions of the parameters discussed in section 6 and appendix B.

While the infectious dose for any set of parameters can be estimated using the equations in section 4, estimating the probability of infection in a given situation requires marginalizing over the probability distribution function associated with each of the parameters. Fortunately, marginalization is made simpler by the fact that *D*_inf_ is proportional to *ρV*_src_/*N*_inf_ in all dose calculations, allowing us to connect *D*_inf_ computed from parameters in Table 1 to *P*_inf_ by introducing the concept of a “nominal infectious dose” 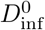. Figure 1 (left) shows the cumulative distribution of *D*_inf_ relative to the nominal value 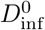 obtained using the parameters in Table 1.

Similarly, the “random *ρV*_src_*N*_inf_” curve in Fig. 1 (right) can be used to estimate the probability of a contagious individual causing infection in the people they interact with, given a nominal infectious dose 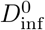 computed with the parameters in Table 1. For instance, using Fig. 1 (right, blue curve) we can estimate that a brief encounter which delivers 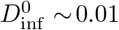 results in *P*_inf_ ~ 2%. This infection probability accounts for parameter variation relative to the nominal values, so no further computation is required.

The infectious dose can be summed over multiple encounters to compute the probability of infection in at least one susceptible individual. For example, if a contagious individual delivers a dose of 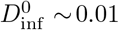 on each of 10 encounters with susceptible individuals, then the total infectious dose is 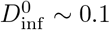 and the probability of transmission to at least one person is *P*_inf_ ~ 6% (again using Fig. 1, right). The distribution of virions via a building’s HVAC system, discussed in section 4.2, provides a similar example of a weak interaction with a large group of people (occupants of rooms connected via the HVAC system).

Note that the final probability of transmission is *not* the sum of the infection probabilities for each susceptible individual because of the correlation between these probabilities (i.e., the viral load of the contagious individual is common to all events). It is exactly this correlation that leads to large outbreaks: if a highly-contagious individual delivers an infectious dose *D*_inf_ ~ 2 to many individuals they will each have a 80% probability of being infected (“random *N*_inf_” curve in Fig. 1, right).

The above example shows that limiting the number of potential transmission events between members of a group, and limiting group size, are both important to minimizing transmission and preventing outbreaks. If only 4 encounters are allowed per person, and the contagious individual only encounters 2 people, then 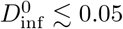 per encounter gives a total 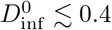 and a 10% probability of transmission.

## 8 Conclusions

The new world of COVID-19 is here and we will all have to learn to live in it. Understanding how to navigate the dangers of this new world will be necessary as we come out of our houses and return to our workplaces. The calculations presented here suggest that keeping the risk of infection low in the workplace may require mitigation techniques and protocols that limit the infectious dose a contagious individual can deliver to their coworkers.

Some broad guidelines which we draw from the above analysis are:

1. Aerosol build-up in closed spaces should be treated with care. Avoiding infection requires good ventilation and/or short exposure times. Generally, office spaces should not be occupied by more than one person. In the early phases of epidemic decay, airflow in shared spaces should be at least 50 m^3^/min ~ 2000 CFM per occupant beyond the first. As the prevalence of COVID-19 in the population decreases this could be reduced as low as 5 m^3^/min ~ 200 CFM per occupant beyond the first (see section 4.1).
2. Recirculation in HVAC systems should be avoided if possible. High quality filters (e.g., MERV 12) should be used in recirculating HVAC systems, and office use should be kept to a minimum to avoid transmission through the air circulation. In the early phases of epidemic decay, airflow should be at least 10 m^3^/min ~ 350 CFM per occupant (averaged over the full HVAC system distribution, see section 4.2).
3. Small or poorly ventilated common spaces where many people spend time (i.e., bathrooms, elevators and stair wells) are of particular concern. Consider increasing airflow and/or adding local air scrubbers to avoid wait times between occupants. For single occupancy shared spaces, 10 m^3^/min ~ 350 CFM should be considered a minimum (see section 4.3).
4. Mask use may help to prevent direct exposure when a minimum of 2 m interpersonal distance cannot be maintained, but is not sufficient to prevent infection in an enclosed space *regardless of the distance between occupants*. Distances less than 1 m remain more dangerous than larger distances due to aerosol leakage around masks, especially in the event of coughing or sneezing. Additional personal protective equipment (PPE) should be used for tasks which require close proximity (see sections 4.4 and 5.1).

## Data Availability

All data used in this work is derived from publicly available sources.

## 9 Acknowledgements

The author would like to thank Lisa Barsotti, Paola Rebusco, Rich Abbott, and Paola Cappellaro for their careful reading and editing of this article. The analysis presented here would not have been possible without the inspiration provided by Dawn Mautner, the information supplied by Scott Kemp and Peter Fisher, and the many hours of quiet time for which I have Lisetta Turini to thank. An early draft of [77] and feedback from its authors were both very helpful and much appreciated. Subsequent feedback from Lisa Brosseau and Dave Boggs has been critical to deepening our literature base and refining our presentation. The author is, as always, very grateful for the computing support provided by The MathWorks, Inc.

## A Outbreaks

Documented outbreaks offer a means for checking the plausibility of our aerosol transmission model. In this section we select a few outbreaks where transmission via aerosols appears likely (e.g., distances were too large for droplets, or the pattern of infection followed the airflow).

Information about the outbreaks is, however, limited, and the viral load of the contagious individual is not known, so these comparisons only provide weak constraints on the model parameters. In particular, reported outbreaks are likely to involve unusually contagious individuals (a.k.a. “super-spreaders”), with *ρ*_0_ ≿ 1000 /nL (roughly 10% of cases), so we use that value in these computations. Since this fixes the value of *ρ*, we use the “random *V*_src_*N*_inf_” curve in Fig. 1 to compute the probability of infection.

### A.1 Guangzhou Restaurant

[88, 89] document a COVID-19 outbreak associated with air flow in a restaurant (Guangzhou, China). Unlike many outbreaks, this one was studied in great detail, including on-site experimental recreation of the airflow and computational fluid dynamics (CFD) modeling, making it an excellent cross-check for our simplified aerosol model [88].

In this outbreak, there was one contagious individual (CI) seated at a table with 9 family members. Air circulation in the restaurant was dominated by wall mounted air conditioning units, one of which essentially defined a well mixed air space that included 2 other families totaling 11 people. The distance between CI and the people infected varied from ~ 50 cm to 4.6 m with 4 of the 5 people who were infected at other tables 2 or more meters from CI [88]. There is no obvious correlation between who was infected and their distance from CI or the direction that CI was facing.

Of the people at the table with CI, 4 of 9 were infected, while 5 of the 11 people at the other 2 tables were infected (45% attack rate in both cases). The aerosol concentration at these three tables was found to be quite similar, both experimentally and in CFD models [88]. Given the common source (i.e., the contagious individual), we use the “random *N*_inf_” curve in Fig. 1 (right, yellow) to estimate the infectious dose required to produce this attack rate as *D*_inf_ ≃ 0.6. In the next paragraph we compare this value with the estimate we get from direct calculation using Eqn. 9.

The recirculating air space defined by the wall mounted AC unit which transported air from CI to other diners who were infected is roughly 60 m^3^ and was found to have *r*_room_ ~ 1 m^3^/min, and roughly 1 air change per hour (*τ*_room_ ~ 60 min). The overlap in time between CI and the other diners in this air space was approximately 60 minutes. This is situation is not in the well ventilated limit, and has *f_a_* ~ 1/4 according to Eqn. 7 with our estimated 20 minute time-constant for settling (see section 6.2).

Assuming a somewhat talkative contagious individual (*r*_src_ ~ 4 nL/min), and given *r*_room_ ~ 1 m^3^/min and *t* ~ 60 min, the other diners were exposed to a *nominal* infectious dose of 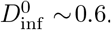. This is a perfect match to the value computed using the attack rate, and indicates that CI’s viral load was likely around *ρ*_0_ at the time of this outbreak. This is not surprising since, as noted above, there is a selection bias when working with noteworthy outbreaks that precludes low values of *ρ*.

The detailed analysis of this outbreak offers a number of other quantitative and qualitative tests of our simplified model of aerosol infection. [88] notes that a table near the CI (“table 17”), but not in the air space defined by the AC unit, had a relatively high aerosol concentration due to leakage from the primary air circulation pattern. There were 5 people at this table, and they overlapped with CI for 18 minutes at the end of CI’s stay. It is unlikely that the infectious dose summed over all 5 diners was larger than 1, since *P*_inf_ ≃ 0.7 for *D*_inf_ = 1, indicating an average *D*_inf_ ≾ 0.2 with 70% confidence (and average *D*_inf_ ≾ 0.6 with 90% confidence). This is consistent with the shorter overlap time and the somewhat reduced aerosol concentration at table 17.

However, a similar analysis applied to the 50 diners who were at more distant tables (e.g., “table 10” in [88]), none of whom were infected, indicates a relatively short aerosol decay time. Since the total infectious dose required to cause at least one infection does not depend on the number of people exposed (i.e., total *D*_inf_ ≾ 1 with 70% confidence, and *D*_inf_ ≾ 3 with 90% confidence), the average infectious dose for these 50 diners must have been less than *D*_inf_ ≾ 0.06 (90% confidence). Assuming an average overlap time with CI of at least 30 minutes, the virion aerosol concentration must have been *at least 5 times less* than the concentration around the 3 tables where the attack rate was high. Using the data in figure S4 of [88], we can see that this requires the aerosol concentration to come to steady-state much more quickly than the tracer gas used in that study. Steady state was likely achieved in less than 10 minutes, and certainly in less than 20 minutes. This agrees with the settling times of 5–10 *μ*m droplet nuclei, as discussed in section 6.2.

Analysis of surveillance videos reveals no close contact, or objects shared, among members of different families. None of the restaurant staff were infected, nor were patrons of other businesses in the building. Simply put, this outbreak is essentially impossible to explain with surface contamination or large droplet driven transmission, and yet easily understood in terms of aerosol transmission.

### A.2 Hunan Coach

[90] documents an outbreak on a long distance coach (Hunan, China). This 45 person coach should have *r*_room_ ≃ 8m^3^/min [11], and we will estimate the ride duration as 2 hours. The contagious individual did not interact with others, so we assign a source rate of *r*_src_ = *r_b_* = 1 nL/min. The resulting nominal dose of 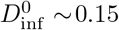 is in reasonable agreement with the fact that 8 of the 45 passengers were infected. The infections were somewhat localized, with the most distant 4.5 meters from the contagious individual, possibly due to incomplete mixing of the air in the bus.

This outbreak also resulted in the infection of a passenger who boarded 30 minutes after the contagious passenger disembarked. This could have been due to surface contamination, but then one must wonder why no passengers on later voyages were infected (since fomites last for days) [76]. More likely it indicates settling and air exchange while the bus was stopped resulted in an order of magnitude reduction in infectious dose. (Outbreaks on vehicles were common in China, possibly due to poor ventilation [11].)

### A.3 Seattle Choir

[91, 92] Of 60 singers 87% infected after singing together for 2.5 hours in a small church (*V*_room_ ~ 600 m^3^). An air-cycle time of 30 minutes gives *r*_room_ ~ 20 m^3^/min, and singing can be approximated by *r*_src_ ~ 30 nL/min. The 150 minute exposure yields an infectious dose of *D*_inf_ ~0.9 (accounting for settling with *f_a_* = 0.4). Sadly, this makes the probability of infection quite high (*P*_inf_ ~ 60%). Even more sadly, this is not the only choir outbreak [93, 94], and this can happen even if distancing and hand-hygiene rules are strictly followed [95]. The least we can do is to learn from their tragic experience. (Like choir practice, aerobic dance classes [96] and call centers [97] are ideal environments for SARS-CoV-2 transmission.)

### A.4 Diamond Princess

[98] concludes from the outbreak on the Diamond Princess cruise ship that long-range airborne transmission is unlikely since SARS-CoV-2 did not pass through the ship’s air conditioning system. Two Okinawa taxi-drivers were, however, infected by passengers during a shore visit. This outbreak is different from the others in that the contagious individual who initiated the outbreak was not involved in the taxi-driver infections. At the time of the shore visit in Okinawa there were roughly 25 pre-symptomatic infected passengers on the Diamond Princess at least a few of whom disembarked.

If we assume that the taxi ride lasted 10 minutes (the port in Naha is close to the main attractions), that the contagious passenger spent the ride talking to another passenger (or coughed once), and that the taxi had the fan on low (*r*_room_ ~ 1 m^3^/min), the driver was exposed to an infectious dose of *D*_inf_ ~ 1 according to Eqn. 9. Using the “random *ρV*_src_*N*_inf_” curve in Fig. 1 since these were secondary infections, we find that the probability of transmission was about 15% for each contagious passenger who disembarked. This probability is relatively insensitive to our assumptions and would only change by a factor of 2 if the dose changed by an order of magnitude (in either direction). If both infections in Naha were caused by the same contagious individual (one on the ride in, one on the ride out) it would be appropriate to use the “random *V*_src_*N*_inf_” probability of 60%.

Fortunately, there were no other COVID-19 infections linked to the Diamond Princess’ stop in Okinawa, indicating that the closed environment and long exposure time of the taxi was likely a key ingredient for transmission. That is, changing the air-flow rate in the above calculation to *r*_room_ ≿ 100 m^3^/min to represent shops and more open spaces would reduce the probability of infection to less than 2%. And, while surface transmission could explain transmission to taxi drivers, it does not explain the absence of transmission to shop keepers or other people with whom the passengers of the Diamond Princess interacted.

### A.5 Hospital Air Sampling

There was no outbreak in the Nebraska Medical Center, but air sampling in and around COVID-19 patient rooms [99] offers a further cross-check of the calculations presented here. Approximately 3 viral RNA copies per liter of air sampled were found in a patient’s room (and in the hallway outside the patient’s room after the door was opened). These rooms have *τ*_room_ ≃ 8 min and *V*_room_ ≃ 30 m^3^, indicating an airflow rate of *r*_room_ ~ 4 m^3^/min [56]. Equation 5 indicates a steady-state concentration of 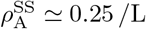. This suggests that either this patient had a very high viral load (*ρ* ~ 10^10^ /mL, which is in the top 3%), or that they were occasionally talking or coughing and had *ρ* ~ *ρ*_0_.

Air sampling data is also available from hospitals and public areas in China. [100] reports quantitative viral deposition rate of roughly 2 /m^2^ per minute in a patient’s room. The area sampled was 3 m from the patient’s bed, so too far for most droplets [14]. If we assume that this is due to slow settling of the larger droplet-nuclei with characteristic a speed of *u*_settle_ ~ 0.1 m/min (see section 6.2), this implies a aerosol concentration of roughly 20 /m^3^. (Oddly, air samplers in patient rooms did not detect concentrations above their detection threshold, but the patient’s bathroom and other areas in the hospital had concentrations near 20 /m^3^).

A concentration of 20 /m^3^ is more than a factor of 100 lower than that reported in Nebraska [99], but still implies p above the median of the distribution function shown in Fig. 2. If we assume an air flow rate in the room of *r*_room_ ~ 4 m^3^/min and viral shedding dominated by breathing (i.e., *V*_src_ ~ *V_b_*), for instance, *ρ* is roughly 10^8^ /mL = *ρ*_0_/10.

**Figure 2:**
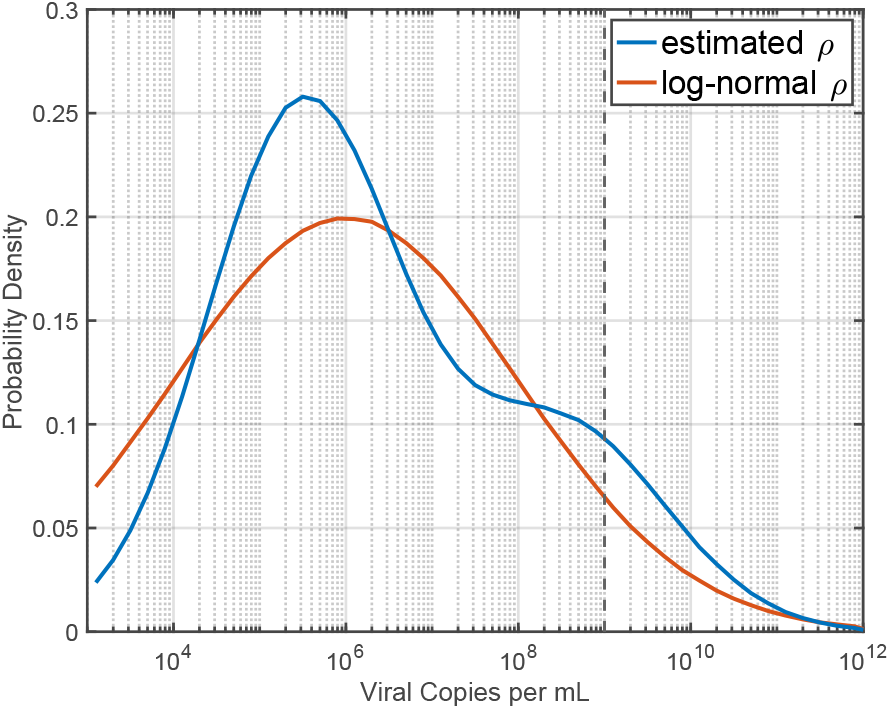
Probability density functions for *ρ*. The estimated distribution used in this work is shown along with a log-normal distribution centered at 10^6^ /mL. The vertical line at 10^9^ /mL indicates our “nominal *ρ*_0_” of 1000 /nL. 10% of the estimated *ρ* distribution lies above this line and 7% of the log-normal distribution lies above it.

## B Parameter Probability Distributions

As described in section 6 several parameters used in this work vary significantly between individuals and events. This appendix describe the details of the probability density functions use for *ρ*, *V*_src_ and *N*_inf_.

Figure 2 shows the distribution for *ρ*. The estimated distribution used in this work is derived from [47] with the low end of the distribution pushed up somewhat to account for the downward trend in viral load after symptom onset. The log-normal distribution centered at 10^6^ /mL with *σ* = 10 is estimated from the linear fit in figure 2 of [49]. These distributions both have a median viral load of 10^6^ /mL and roughly 10% of cases above 10^9^ /mL. We have computed *P*_inf_ as in Fig. 1 for both distributions and they give quantitatively similar, and qualitatively identical, results.

To account for individual variability, we use a log-normal distribution for *N*_inf_ which is centered around 1000 and has a width of a factor of 2. This makes the 95% confidence interval 250–4000 which is in reasonable agreement with [42] and [31].

Similarly, an order of magnitude variation among individuals and events is also expected in droplet and aerosol production [80, 25]. For this we also use a log-normal distribution centered around the *V*_src_ values given in Table 1 with a width of a factor of 3.

